# Protecting our health care workers while protecting our communities during the COVID-19 pandemic: a comparison of approaches and early outcomes in two Italian regions, 2020

**DOI:** 10.1101/2020.04.10.20060707

**Authors:** Nancy Binkin, Federica Michieletto, Stefania Salmaso, Francesca Russo

## Abstract

**Introduction:** Italy, which has been hard-hit by the COVID-19 pandemic, has an overriding national strategy, but its 21 regions have adapted their response based on the organization of their curative and public health services. In this paper, we compare short-term outcomes for two northern Italian regions which had almost simultaneous initial outbreaks: Lombardy, which had a patient-centered approach that relied on primary care physicians and hospital care, and Veneto, which focused on community-based diagnosis and care.

**Methods:** We used numerator and denominator data from public Italian government sources to calculate reported rates of COVID-19 testing/1000, COVID-19 cases/100,000 overall and for health care workers (HCWs) and non-HCWs, deaths per 100,000, and the percent of cases admitted to hospitals and ICUs for February 24-April 1, 2020.

**Results:** As of April 1, 2020, Lombardy experienced 44,733 cases and 7,539 deaths; for Veneto, the corresponding values were 9,625 and 499. The cumulative case rate was 445/100,000 for Lombardy and 196/100,000 for Veneto, a 2.3-fold difference. Mortality rates were 7.5 times higher in Lombardy than in Veneto (75/100,000 and 10/100,000, respectively). Cumulative rates of testing were nearly twice as high in Veneto and were 2.7 times higher in the first week of the epidemic. In Lombardy, 51.5% of patients were admitted, including, 5.2% to intensive care units; for Veneto, the corresponding figures were 25.1% and 4.3%, respectively. HCWs account for 14.3% of all cases in Lombardy compared with 4.4% in Veneto. In Lombardy, the rate among HCWs was 19.1 times higher than in the general population (6,924/100,000 versus 362/100,000), while in Veneto it was 3.9 times higher (676/100,000 versus 172/100,000).

**Discussion:** The community-based approach in Veneto appears to be associated with substantially reduced rates of cases, hospitalizations, deaths, and infection in HCWs compared with the patient-centered approach in Lombardy. Our findings suggest that the impact of COVID-19 can be reduced through strong and aggressive public health efforts to confirm and isolate initial cases and contacts in a timely way and to minimize unnecessary contact between HCWs and cases through home-based testing and pro-active home follow-up.

## Introduction

Italy has been hard-hit by the COVID-19 pandemic, which began simultaneously in two northern Italian regions, Lombardy and Veneto, during the third week of February 2020. By April 1, 2020, Italy had experienced over 110,000 cases and more than 13,000 deaths (1).

In Italy, the National Health Service (NHS) provides preventive services, primary and specialist care, and hospital care free of charge to all citizens and legal residents (2). Each resident is registered with a general practitioner (GP). The GPs serve as primary providers and gatekeepers for specialty care and are paid by the government on a per capita basis, irrespective of the health status of their patients. Both public and private hospitals receive Diagnosis Related Group (DRG) compensation from the government. Each of its 21 administrative areas (19 regions and two Autonomous Provinces) is responsible for local organization of its health services.

Italy developed an overriding national strategy for COVID-19 response and implemented a national lockdown on March 12, 2020. Within these broad guidelines, each region developed a response plan that took into account the strengths and weaknesses of its curative and public health services (3, 4).

With the aging of the Italian population and increasing health care costs, funding and personnel of regional public health programs have declined over the last two decades. The magnitude of the decline has varied between regions and increasing divergence across the regions has been observed in their relative emphasis on public health and curative services. In collaboration with the private sector, some regions, including Lombardy, have created an extensive network of curative services and hospitals but have decreased funding for public health field activities and public laboratories (4). Others, such as Veneto, have continued to support a strong public health network with community outreach. Organizational changes have occurred over time that reflect the changing emphasis on curative services. In some regions, including Lombardy, public health services and hospitals have separate management structures, while in others, such as Veneto, most or all of the hospitals are managed by local health units that also provide preventive services. The response to the pandemic in the two regions reflects these differences.

The organization of health services in Lombardy and Veneto, along with other factors, influenced the approaches initially taken in the critical early weeks of the epidemic. Lombardy employed a patient-centered approach relying primarily on its comprehensive curative services network to deal with the epidemic, while Veneto implemented a broad community-based strategy that relied on its more robust public health network and local integration of services (3).

In this observational study, we present the key elements of Veneto’s community-based approach and compare rates of case-finding, number of cases, hospitalization, deaths, and health care worker (HCW) and non-HCW infections between Lombardy and Veneto.

### Background

#### Demographic and health system characteristics of Lombardy and Veneto

Lombardy has a higher population density than Veneto (420/km2 versus 270/km2) and a higher gross domestic product (GDP; 35, 234 versus 30,445€ in 2017) (5). However, indicators are virtually identical for all 11 Organization for Economic Cooperation and Development (OECD) indicators of well-being, including health (6), as are average age (45.9 versus 45.4 years) and life expectancy (84.0 versus 83.9 years) (7). Both regions have international airports, are heavily involved in international commerce, and are tourist destinations and thus are likely to have similar risks of exposure to imported pathogens.

While the number of acute hospital beds per 1000 is virtually identical (3.05 in Lombardy versus 3.01 in Veneto), the number of adults per GP is slightly higher in Lombardy (1,400) than in Veneto (1,342), physicians and dentists/1000 population is slightly lower in Lombardy (1.3 versus 1.6) (8). Per capita health expenditures are also similar (7). In the public health domain, however, differences are far greater. In Lombardy, there are three public health laboratories (approximately 1 per 3 million population) (4) while in Veneto, there are 10 (approximately 1 per 0.5 million). There are 8 public health prevention departments in Lombardy (1 per 1.2 million) compared with 9 in Veneto (1 per 0.5 million) (9). Home-based care is more common in Veneto than Lombardy, as evidenced by participation in Assistenza Domiciliare Integrata (ADI; integrated home assistance) that provides in-home services to the elderly, the disabled, and those with chronic conditions. In 2017, the most recent year for which data are available, the program served 3.5/100,000 in Veneto versus 1.4/100,000 in Lombardy (8).

#### COVID 19 approach in Lombardy

The first case in Lombardy was identified on February 20 in Codogno (10), a town of 15,000, which was placed in lockdown by the national government on February 24, 2020. Over a 7-day period, from February 24 to March 2, the number of cases in Lombardy expanded 6.5-fold, from 166 to 1077 (1.6 to 12/100,000 population) (11). Early on, three foci rapidly emerged (10).

Initial efforts by the Regional Health System focused on three primary objectives, including collection of data to understand the epidemiology and conduct modeling, to increase diagnostic capacity, and to promote-hospital based assistance for cases (10), and efforts were made to also introduce isolation and contact tracing. Additionally, the existing strong regional ICU network was potentiated (12). Guidelines were issued for GPs regarding diagnosis, testing, and referral to hospitals (13). Testing was initially focused on those with symptoms as per national policy, and contact tracing and home-based testing, care, and follow-up efforts were hampered by the rapid explosion of in the number of cases (4).

In the resulting patient-centered approach, physicians, ambulatory clinics, and emergency rooms served as the front line during the COVID pandemic (3, 4, 10, 16). In the absence of other options, patients were sent to the hospital, overwhelming the existing human resources and beds and essentially diluting the quality of care (16). Patients were referred to the infectious disease services in 17 hospitals throughout the region (13). Convalescent centers for those not needing acute care but who needed continued monitoring were not available until weeks after the beginning of the epidemic.

#### COVID 19 approach in Veneto

In Veneto, the first cases occurred in Vò Euganeo, a rural village of 3000 people, on February 20, 2020. Like Codogno, the area was put on lockdown on February 24 and extensive testing of residents was performed. Between February 24 and March 2, the cases in Veneto increased 8.5-fold, from 32 to 271 (0.6/100,000 to 5/100,000) (11).

Health authorities in Veneto identified hospitals and convalescent centers that would care for COVID-19 cases, doubled the region’s ICU capacity and obtained an adequate number of ventilators. They gradually moved non-COVID-19 patients out of the designated COVID-19 hospitals, generally to smaller community hospitals set aside for non-COVID-19 patients. In addition to strengthening the ability to care for patients, however, enhanced public health measures were also developed and implemented. An articulated community-based strategy was implemented. Key elements are presented in the Box, but they included extensive contact tracing, rapid testing of both cases and an extended network of contacts, supervised quarantine and isolation, minimization of contact between HCW and the public, and informatics systems for rapid communication on case diagnosis and management and for monitoring bed availability. All non-essential public health activities were promptly put on hold, and a force of more than 750 public health workers throughout the region was mobilized.

## Methods

### Data sources

We obtained data on the region-specific numbers of tests performed, cases, deaths, hospital and ICU admissions, and patients in home care for February 24 through April 1, 2020, from archived official daily bulletins of the Protezione Civile (11), the branch of the government that manages national emergencies. Data on cumulative cases of COVID-19 infection in HCWs for Lombardy and Veneto were obtained from region-specific data published April 3, 2020, by the Istituto Superiore di Sanità (14).

Because the population of Lombardy is more than twice that of Veneto (10.1 versus 4.9 million), and correspondingly, the number of HCWs also differs, the data on testing, cases, deaths in the overall population, and cases in HCWs are presented as rates. Regional population denominator data were obtained from census projections (7). Data on the number of HCWs by region was taken from the most recent yearbook of the Statistics Office of the Ministry of Health (8), which examines the number of publicly employed HCWs, who comprise the vast majority of health sector employees. For analyses comparing rates among HCWs and non-HCWs, the denominator of non-HCWs was obtained by subtracting the number of publicly employed HCWs from the overall population in each region.

Cases were defined as persons who were COVID-positive based on PCR testing, according to the national case definition (15).

## Results

### Testing

As of April 1, 2020, the number of tests performed per 1000 residents in Veneto was nearly twice that of Lombardy (23.0/1000 residents in Veneto; >112,000 tests vs. 12.1/1000 in Lombardy). The difference was even greater in the first critical week, when there were 2.7 times as many tests done in Veneto.

### Cases and deaths/1000 population

Trends in cases and deaths for Lombardy and Veneto are shown in Figure 1. Within a week of the first cases, case notifications in both regions diverged, and mortality began to diverge one week later. As of April 1, 2020, there were 44,733 cumulative cases in Lombardy and 9,625 in Veneto; the numbers of deaths were 7,593 and 499, respectively. The cumulative case rate was 445/100,000 for Lombardy and 196/100,000 for Veneto, a 2.3-fold difference. Mortality rates were 75/100,000 and 10/100,000, respectively, a 7.5-fold difference. The death-to-case ratio was 3.3 times higher in Lombardy than in Veneto (17% versus 5%).

**Figure 1.**
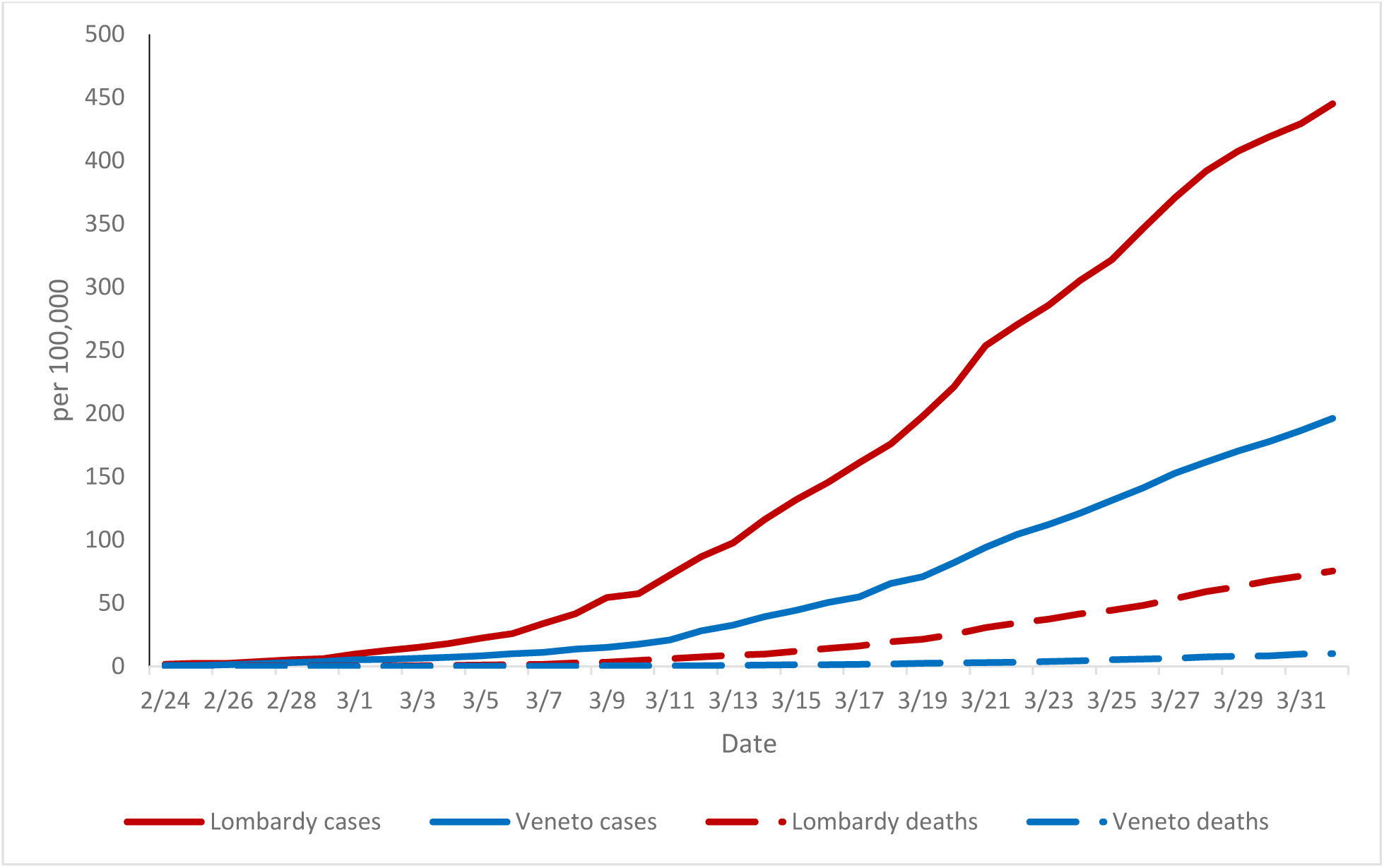
COVID 19 cases and deaths per 100,000, Lombardy and Veneto, 2/24-4/1/2020

### Disposition at diagnosis

As of April 1, 2020, 51.5% of COVID-19 cases in Lombardy had been admitted to the hospital, including 5.2% to ICUs and 46.3% to acute care. Veneto admitted 25.1% of COVID-19 cases (4.3% to ICUs and 20.9% to acute care). In Lombardy, the remaining 48.5% were placed in home isolation, compared with 74.9% in Veneto.

### Infection in HCWs

As of March 30, 2020, 14.3% of all COVID-19 cases in Lombardy were among HCWs, compared with 4.4% in Veneto. Figure 2 shows the difference in the rates of COVID-19 infection in HCWs compared with non-HCW. In Lombardy, the rate among HCWs was 19.1 times higher than in the rest of the population versus 3.9 times higher in Veneto.

**Figure 2.**
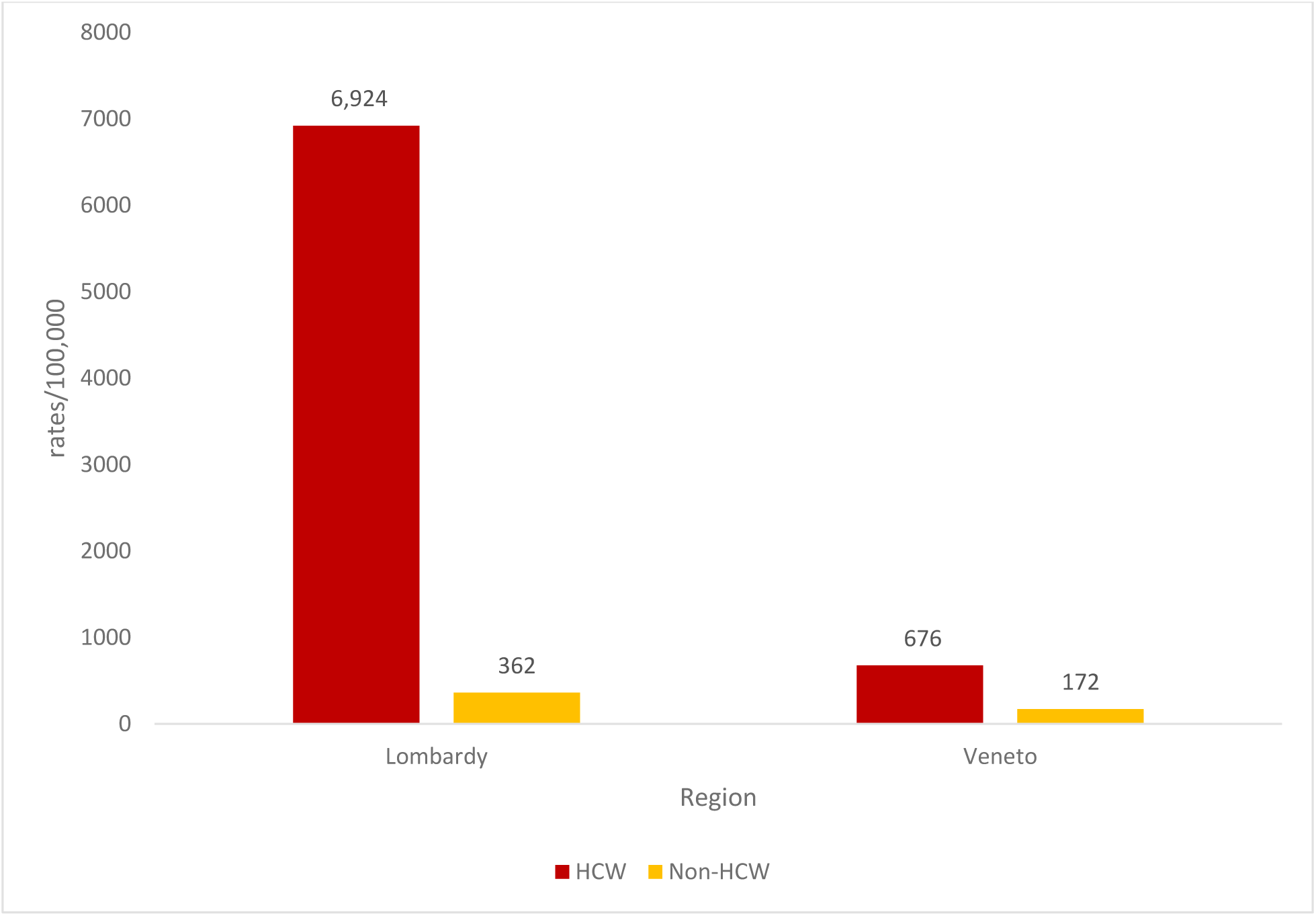
Comparison of cumulative rates/100,000 of COVID-19 infection among HCWs and non-HCWs, Lombardy and Veneto, 3/30/2020

## Discussion

While it is too early to judge the ultimate success of the response, Veneto’s community-based approach appears to have reduced a wide range of adverse outcomes during the initial weeks of the epidemic. In Veneto, case rates, death rates, and health care worker infections were considerably lower than in Lombardy, despite the valiant and courageous efforts of the many dedicated providers (12, 17) to activate and potentiate their extensive patient-centered care network.

Differences in population density and social factors, as well as the greater initial number of cases in Lombardy and greater numbers of initial foci, may have played a role in the observed differences in outcomes (3). However, health care system organization and the strength of the public health infrastructure ultimately appear to have played an important role in the differences in outcomes observed to date between Lombardy and Veneto (3, 4, 16, 17). As stated by clinicians from one of the most heavily affected hospitals in Lombardy, “Western health care systems have been built around the concept of *patient-centered care*, but an epidemic requires a change of perspective toward a concept of *community-centered care*.” (17)

Veneto’s greater integration of its public health and hospital services at local level and strong public health infrastructure potentiated the implementation of a community-based approach. This approach was based on sound epidemiologic principles of aggressive testing, contact tracing, and limiting contact with health care settings wherever possible through mobile diagnostic teams and careful home follow up and was facilitated by rapid communication through an informatics system that tied together the laboratory, GPs, and the local public health units.

Early and aggressive testing to diagnose COVID-19 in cases and their contacts likely played a critical role in Veneto’s trajectory and outcomes. In the first week, the rate of testing/1000 population in Veneto was 2.7 times higher than Lombardy, which may have been critical in limiting further spread. Furthermore, the broader definitions of contacts to include extended family, work, and more casual contacts and the subsequent testing and isolation of these individuals also was likely to have been a major contributor to early reduction in spread.

The approach of protecting GPs in the community by emphasizing telephonic rather than in-person visits and using a mobile public health team to obtain specimens and evaluate the condition of patients on home monitoring appears to have protected practitioners in Veneto and may have limited their role in amplifying community spread. GPs’ offices concentrate older persons and those with chronic health conditions. Also, GPs in Italy often make home visits to patients with limited mobility. While it is not possible to demonstrate transmission from GPs to their patients, it is clear that many GPs in Lombardy were infected. Based on data from the registry of COVID-19 related deaths among physicians maintained by the national association of physicians and reported in an international registry (18), 17 physicians listed as GPs in the Lombardy region have died as of April 1, 2020, compared with zero in Veneto.

Nosocomial transmission appears to have played a role in transmission in Lombardy (16). Efforts to keep COVID-19 patients away from health care facilities during diagnosis and provide home follow up for patients wherever possible appear to have reduced HCW risk of infection. In Veneto, where only a quarter of diagnosed cases were hospitalized, less than 5% of cases were in HCWs. In Lombardy, where over half of diagnosed cases were hospitalized, the corresponding value was 14%. The rate of infection in HCWs in Veneto was almost four times higher than the remainder of the population, clearly suggesting that nosocomial transmission was occurring, but was far lower than in Lombardy, where HCWs were 19 times more likely to be infected than the general population.

The explosive nature of the epidemic in Lombardy rapidly overwhelmed its initial efforts to maintain separate COVID-19 facilities, and it became necessary to admit cases to hospitals containing non-COVID patients. Having COVID-19-dedicated facilities in Veneto in the first weeks of the epidemic, which involved moving non-COVID patients to other facilities to allow creation of COVID-19-specific hospitals and convalescent centers, may also have contributed to limiting health care worker infection and the spread in the community to vulnerable non-COVID patients and their visitors. This strategy also allowed exposed HCWs to more efficiently and effectively use scarce personal protective equipment (PPE). Furthermore, heavy environmental contamination has been documented in locations where care is provided for COVID-19 patients (19) and maintaining adequate protection to prevent transmission to uninfected patients may be particularly difficult in overwhelmed hospitals that have limited supplies of PPE.

This study has several limitations. It is an observational study based on the experience of only two regions, and because of its observational nature, it is difficult to draw indisputable causal relationships. Lombardy experienced a more rapid initial explosion of cases, and had the onset been slower, there might have been time to organize and implement more public health measures to reduce transmission in the community. Additionally, this study represents a point in time, with Italy now only beginning to experience a decline in new cases. As cases continue to occur, the presumptive impact of the Veneto approach may lessen as the public health system and hospitals become increasingly strained.

In terms of methodological limitations, case and death reporting may have differed between the two regions. Veneto, with its real-time integrated information system, may have had more complete and accurate case reporting. However, if cases and deaths were more likely to be notified than in Veneto than in Lombardy the effect would be to underestimate rates for Lombardy and increase, rather than decrease observed differences in the case and death rates between the two regions. Differences in HCW testing could have also affected our results. HCWs in Veneto who have been in close contact with cases but do not have symptoms are tested three times over fourteen days as part of the policy to keep them on the job. This testing policy in Veneto would likely lead to the detection of more HCW cases compared with Lombardy, but the effect would be in the direction of increasing the observed difference in HCW rates of infection between Veneto and Lombardy. Finally, our denominators for HCWs did not include those employed privately. However, the vast majority of all HCWs in the country are employed by the government and would be unlikely to affect the magnitude of the differences observed between the two regions.

Currently, the applicability of the community approach may be limited to countries where public health and curative services are integrated. In other countries such as the United States, where health care is privatized, and integration with public health services is limited, it nonetheless may be possible to implement most of the major components in specific settings. One such setting may be Kaiser Permanente, which covers an estimated population of 12.2 million and integrates prevention and treatment, has its own laboratories and hospitals, has a tradition of community-based care, maintains a sophisticated information system, and often has good working relationships with local health departments that are responsible for contact tracing. However, even in settings where full implementation is not possible, it may be feasible to identify human and financial resources to increase non-facility-based screening, contact tracing, and home follow-up activities rather than providing expensive and labor-intense hospital-based care, which appears to result in poorer patient and HCW outcomes. In preparation for the end of the extensive lock-down and to avoid additional waves, every effort should be made to strengthen health department capacity for extensive contact tracing and isolation, community monitoring of patients, and communication between public health and health care providers and to establish robust informatics systems that allow for real-time information transfer among the many entities involved in controlling community spread. Finally, markers and indicators of the public health performance are needed to measure the public health capacity to adequately respond to such threats.

## Data Availability

Data available from public sources: see manuscript for links

## Acknowledgments

The authors wish to thank Dr. Sandra Berrios-Torres for her careful review and comments, Judy Berman Seibert for her editorial assistance, and Karen Heskett for her invaluable bibliographic support.

## Box Key elements, Veneto Community-Based Strategy

The key elements of the Veneto strategy are as follows:

- Extensive testing of symptomatic and asymptomatic cases with rapid laboratory turnaround
- Broad contact tracing around test-positive cases, including extended family, work, and more casual contacts (e.g., at meetings >15 minutes)
- Self-quarantine of cases and suspected cases with daily telephone monitoring to assess clinical status
- Dissemination of detailed and practical guidelines on home isolation to protect other household members
- Minimization of contacts with physicians and other HCWs through:
  - Telephone rather than in-person visits wherever possible by GPs
  - Home testing wherever possible and home visits by a specially dedicated team to assess changes in clinical status as needed
  - Limiting hospital admissions to persons requiring oxygen or who have other major health issues
  - Cohorting of hospitals as COVID-19 or non-COVID-19
  - Creating COVID-19 convalescent hospitals for patients no longer requiring acute care
  - Testing of exposed HCWs three times every 14 days and exclusion of those who have symptoms or have positive tests.
- Extensive informatics systems providing real-time testing results to local health units responsible for contact investigation, results of daily call center findings, and a separate system for monitoring of hospital and ICU bed availability.

## Bibliography

1. Redazione Online. Coronavirus in Italia, 110.574 casi positivi e 13.155 morti. Il bollettino del 1° aprile. Corriere della Sera [Internet]. 2020 Apr 1 [cited 2020 Apr 7]; Available from: https://www.corriere.it/salute/malattie_infettive/20_aprile_01/coronavirus-italia-110574-casi-positivi-13155-morti-bollettino-1-aprile-766df6fc-742c-11ea-b181-d5820c4838fa.shtml

2. Donatini A. The Italian health care system [Internet]. The Commonwealth Fund: International Health Care System Profiles. [cited 2020 Apr 7]. Available from: https://international.commonwealthfund.org/countries/italy/

3. Pisano GP, Sadun R, Zanini M. Lessons from Italy’s Response to Coronavirus. Harvard Business Review [Internet]. 2020 Mar 27 [cited 2020 Apr 7]; Available from: https://hbr.org/2020/03/lessons-from-italys-response-to-coronavirus

4. Carra L. Vittorio Carreri: l’epidemia si combatte anche sul territorio [Internet]. Scienza in rete. 2020 [cited 2020 Apr 7]. Available from: https://www.scienzainrete.it/articolo/vittorio-carreri-l%E2%80%99epidemia-si-combatte-anche-sul-territorio/luca-carra/2020-03-26

5. Istituto Nazionale di Statistica. Noi Italia [We Italy] [Internet]. I.Stat. [cited 2020 Apr 7]. Available from: http://noi-italia.istat.it/index.php?id=3&L=1

6. OECD Regional Well-Being - “A closer measure of life” [Internet]. OECD Regional Well-Being. [cited 2020 Apr 7]. Available from: https://oecdregionalwellbeing.org

7. Istituto Nazionale di Statistica. Indicatori demografici [Demographic indicators] [Internet]. I.Stat. [cited 2020 Apr 7]. Available from: http://dati.istat.it/Index.aspx?DataSetCode=DCIS_INDDEMOG1

8. Ministero della Salute, Ufficio di Statistica. Annuario statistico del servizio sanitario nazionale. [Internet]. Rome, Italy: Direzione Generale della Digitalizzazione del Sistema Informativo Sanitario e della Statistica Ufficio di Statistica; 2017 [cited 2020 Apr 7] p. 117. Available from: http://www.salute.gov.it/imgs/C_17_pubblicazioni_2879_allegato.pdf

9. Istituto Nazionale di statistica. Health for All Health for All – Italia. Sistema informativo territoriale su sanità e salute. Versione di dicembre 2019 {Health for all-Italy. Regional information system on the health system and health. Version from December 2019] [Internet] I.Stat [cited 2020 Apr 7]. Available from https://www.istat.it/it/archivio/14562

10. Tirani M, Rovida F, DeMicheli M, Pletti A et al. The early phase of the COVID-19 outbreak in Lombardy, Italy. https://arxiv.org/abs/2003.09320v1

11. Rosini U. dpc-covid19-ita-regioni.csv [Internet]. GitHub. 2020 [cited 2020 Apr 7]. Available from: https://github.com/pcm-dpc/COVID-19

12. Grasselli G, Pesenti A, Cecconi M. Critical Care Utilization for the COVID-19 Outbreak in Lombardy, Italy: Early Experience and Forecast During an Emergency Response. JAMA [Internet]. 2020 Mar 13 [cited 2020 Apr 7]; Available from: https://jamanetwork.com/journals/jama/fullarticle/2763188

13. Regione Lombardia. News. Coronavirus, assessore Gallera: emanate direttive a medici medicina generale e ospedali per presa in carico ‘casi sospetti. [Regional website][internet] 2020 Jan 28 (cited 2020 Apr 7]; Available from: https://www.regione.lombardia.it/wps/portal/istituzionale/HP/lombardia-notizie/DettaglioNews/2020/20-26/coronavirus-gismondo-no-allarmismi

14. Istituto Superiore di Sanità. Epidemia COVID-19: Appendice al Bollettino con il dettaglio regionale [Internet]. Istituto Superiore di Sanità; 2020 Mar p. 54. Available from: https://www.epicentro.iss.it/coronavirus/bollettino/Bolletino-sorveglianza-integrata-COVID-19_30-marzo-2020_appendix.pdf

15. D’Amario C. Circolare del Ministero della Salute 9 Marzo: COVID-19. Aggiornamento della definizione di caso [COVID-19. Updating the case definition] [Internet]. Rome, Italy: Ministero della Salute; Ufficio 5 prevenzione della malattie transmissibili e profilassi internazionale; 2020 Mar [cited 2020 Apr 7] p. 12. Available from: http://www.trovanorme.salute.gov.it/norme/renderNormsanPdf?anno=2020&codLeg=73669&parte=1%20&serie=null

16. Capelli A. Gli errori della Lombardia - Quotidiano Sanità [The mistakes of Lombardy]. Quotidiano on line di informazione sanitaria [Online newspaper of health information] [Internet]. 2020 Apr 6 [cited 2020 Apr 7]; Available from: http://www.quotidianosanita.it/lombardia/articolo.php?articolo_id=83627

17. Nacoti M, Ciocca A, Giupponi A, Brambillasca P, Lussana F, et al. At the epicenter of the Covid- 19 pandemic and humanitarian crises in Italy: changing perspectives on preparation and mitigation. NEJM Catal. 2020; (published online March 21.) https://catalyst.nejm.org/doi/full/10.1056/CAT.20.0080

18. In Memoriam: Healthcare Workers Who Have Died of COVID-19 [Internet]. Medscape. 2020 [cited 2020 Apr 7]. Available from: http://www.medscape.com/viewarticle/927976

19. Santarpia JL, Rivera DN, Herrera V, Morwitzer MJ, Creager H, Santarpia GW, et al. Transmission Potential of SARS-CoV-2 in Viral Shedding Observed at the University of Nebraska Medical Center [Internet]. Infectious Diseases (except HIV/AIDS); 2020 Mar [cited 2020 Apr 7]. Available from: http://medrxiv.org/lookup/doi/10.1101/2020.03.23.20039446

